# Exploratory Study on Blood and Urine Transcriptomes for Identifying miRNA Biomarkers in Pediatric Febrile Bacterial and Viral Infections

**DOI:** 10.1101/2025.03.28.25324176

**Authors:** Tatjana Kiseļova, Baiba Vilne, Gunda Zvīgule-Neidere, Dace Gardovska

## Abstract

There is a lack of fast, reliable and non-invasive methods for distinguishing between bacterial and viral infections in modern clinical practice. It negatively impacts the effectiveness of treatment and increases the risk of unnecessary antibiotic administration and thus antibiotic resistance spread. Using microRNA biomarkers in urine may become the solution to the problems listed. miRNAs in urine are relatively stable and can provide an indirect yet specific insight into systemic infections, offering a promising diagnostic tool. In this pilot study, urine small RNA and blood full transcriptome sequencing data were analyzed in children with bacterial infections (n=7), viral infections (n=7), and controls (n=8). The objectives were to identify exploratory urinary miRNA diagnostic signatures and attempt to indirectly correlate differentially expressed (DE) urinary miRNAs with DE transcripts in blood. Using LASSO regularized logistic regression and ANOVA, miRNA biomarker candidates were prioritized. Cross-compartment miRNA:mRNA interactions were putatively evaluated using correlation analysis and target prediction. LASSO regularized regression identified a 5-miRNA signature yielding an apparent internal AUC score of 0.981, though wide confidence intervals reflect the exploratory nature of this small cohort. The prioritized miRNAs demonstrated a descriptive capacity to cluster the patient groups. Correlation analysis and target prediction highlighted parallel molecular patterns across biological fluids, mapping to key immune processes such as cytokine-cytokine receptor interactions and T-cell activation. Our pilot study suggests that prioritized urine miRNA biomarkers reflect host infectious etiology and merit further investigation. While internal performance metrics require validation in larger, independent cohorts, the non-invasive approach represents a promising step toward targeted, timely treatment and the reduction of antibiotic resistance.

## Background

Fever in children is one of the most common reasons for doctor visits and emergency medical assistance. While it is often caused by self-limiting viral infections that resolve with symptomatic treatment, it can be the first sign of serious bacterial infections.^1^ An accurate and timely diagnosis of infection etiology is critical for effective treatment, as empirical antibiotic administration in suspected bacterial cases is common due to the lack of reliable diagnostic tests. This approach, however, poses risks, including unwanted side effects and spread of antibiotic resistance, which remains a significant challenge, especially in hospital settings.^1,2^ Rapid diagnostics of infection etiology are of vital importance when it comes to improving the efficacy of treatment.^3^ For instance, in case of bacterial sepsis, every hour without antibiotics increases morbidity by 8%.^4^ This challenge is further compounded by the continuous evolution of viral screening platforms and the variable cellular response profiles observed across different pediatric cohorts.^5,6^

Differentiating between bacterial and viral infections based solely on symptoms is challenging due to overlapping clinical presentations.^7,8^ However, on the molecular level, bacterial and viral infections induce distinct signal pathways. Viral pathogens preferentially stimulate intracellular pattern recognition receptors, such as Toll-like receptors (TLR) 3, 7, and 8, alongside RIG-I-like receptors, initiating cascades that heavily depend on interferon (IFN) regulatory factors to drive type I and type III interferon synthesis and the upregulation of viral-specific host markers like MxA, IP-10, and TRAIL. However, bacterial infections predominantly engage surface-bound receptors such as TLR2 and TLR4, activating MyD88-dependent pathways that orchestrate the release of classical pro-inflammatory cytokines (e.g., IL-6, IL-8) and downstream acute-phase reactants including C-reactive protein (CRP) and procalcitonin (PCT), alongside altered expression of cell-surface integrins and complement regulators.^9^ This also implies that host gene expression varies depending on etiology of the infection. It has been proven that the etiology of infections in children caused by viruses, gram-negative bacteria (*Escherichia coli*) or gram-positive bacteria (*Staphylococcus aureus* or *Streptococcus pneumoniae*) can be determined solely based on gene expression profile in blood.^10^ Another study confirms that during influenza A H1N1/09 infection increased expression of inflammatory pathway genes has been observed as well as reduced expression of adaptive immune pathway genes.^11^ Two-transcript signature in blood has been shown to be sufficient to distinguish between bacterial and viral infections in febrile children with high sensitivity and specificity.^12^ A recent review covering studies conducted over the past 15 years highlights the need for further research to realize the full diagnostic potential of transcriptomics.^13^ Signatures discovered in adult patients have been shown to have a decrease in performance in pediatric cohorts which is especially prominent in patients 12 years of age and younger.^14^

Advancement of diagnostic methods through novel technologies is vital to improve treatment precision. Micro (mi)-RNAs are attractive targets for the development of diagnostic methods due to their involvement in gene regulation and high stability in bodily fluids.^15^ miRNAs are small (19 to 25 nucleotides) single-stranded non-coding RNA molecules that regulate post-transcriptional gene silencing. More than 60% of human protein-coding genes contain at least one conserved miRNA binding site.^16^ The role of miRNAs in immune system regulation is being actively studied:^17^ miR-146, for example, reduces excessive inflammation in bacterial infections by downregulating cellular lipopolysaccharide sensitivity, ^18^ while miR-155 influences adaptive immune responses to viral infections.^19^ Studies on miRNA in blood have shown their potential to distinguish between acute respiratory bacterial and viral infections in adults.^20^ The use of urinary transcriptome for diagnostic purposes has been extensively evaluated for diseases related to the urinary tract: various renal disorders,^21,22^ prostate and bladder cancer,^23,24^ as well as urinary tract infections, including in pediatric cohorts.^25^ Beyond these applications, urinary miRNA has shown potential to identify systemic conditions, such as chronic inflammatory diseases in children like atopic dermatitis.^26^ The use of urine in pediatric care and diagnostics is particularly promising because it can be collected non-invasively, unlike blood, which can be difficult to collect in sufficient quantities for diagnostic procedures.

This exploratory pilot study investigates the diagnostic potential of urine miRNA signatures to distinguish bacterial and viral infections in pediatric patients. Using small RNA sequencing data, we identified a miRNA profile capable of differentiating these infections with feature selection methods: penalized logistic regression and ANOVA. Additionally, we evaluated these findings in parallel with a whole blood differential expression (DE) mRNA analysis to examine how localized urinary miRNA alterations correlate with systemic host immune dynamics across distinct biological fluids. This multi-omics analysis is utilized as an exploratory, hypothesis-generating approach to discover synchronized molecular pathways between the two fluids, offering novel insights into systemic-to-renal biological processes during acute infection. These results contribute to the emerging field of non-invasive diagnostic tools and highlight the utility of urine miRNAs in pediatric infection diagnostics.

## Materials and Methods

### RNA extraction and sequencing

Blood samples were collected and preserved in PAXgene Blood RNA Tubes (PreAnalytiX), while urine samples were collected in Urine Collection and Preservation Tubes (Norgen Biotek), according to the manufacturer’s specifications. RNA extraction was performed using the PAXgene 96 Blood RNA Kit for blood samples and the Urine Total RNA Purification Maxi Kit for urine samples, adhering to the manufacturer’s protocol. For blood samples, whole transcriptome paired-end sequencing was conducted with 100 million reads per sample and a fragment length of 150 bp. Library preparation used the MGIEasy RNA Directional Library Prep Set, and sequencing was performed on the MGISEQ-2000 platform with the MGISEQ-2000RS High-Throughput Sequencing Set, following the manufacturer’s recommendations. Urine samples were subjected to single end small RNA sequencing with up to 50 million reads per sample and a fragment length of 50 bp. Library preparation was performed using the MGIEasy Small RNA Library Prep Kit, and sequencing was carried out on the DNBSEQ-G400 platform using the DNBSEQ-G400RS High-Throughput Sequencing Set (Small RNA). Before sequencing, ribosomal RNA was removed from all samples using the MGIEasy rRNA Depletion Kit, according to the manufacturer’s protocol.

### RNA data pre-processing

Quality control of raw sequencing reads was performed using *FastQC* v0.11.9^27^ with *MultiQC* v1.14^28^ for visualisation.

Sequence alignment was performed to human genome build GRCh38.p13.^29,30^ Mature and hairpin reference miRNA sequences were downloaded from *miRBase* (mirbase.org, release 22.1).^31^ Adapter trimming, removing short fragments (<18 nt according to research data on miRNA size),^32^ mapping, and miRNA quantification were carried out using *miRDeep2* v0.1.2.^33^ Since the *miRDeep2* algorithm counts both mature miRNAs and its precursors, those were merged to avoid inconsistencies, and the median counts were calculated.

*STAR* v2.7.8a^34^ with default parameters for paired-end sequencing data was used for full transcriptome sequences mapping and adapter clipping. Quality trimming was omitted since it can potentially worsen mapping results.^35^ Quality control of mapping was performed using *Qualimap* v2.3.^36^ Samples with a very small proportion of mapped sequences (less than 1%) were excluded from the downstream analysis. The mapped .*bam* files from the multiple sequencing lanes of one sample were merged with *Samtools* v1.10.^37^ Gene expression counts matrix was produced using R statistical software v4.3.2^38^ package *featureCounts*.^39^

### Multivariate statistical analysis

After removing miRNAs with consistently low counts (median <10) count matrix was normalized using the TMM method as implemented in the *edgeR* package.^40^

Differential expression analysis for miRNA data was performed by fitting a generalized linear model with Bayesian empirical statistics to smooth the standard errors as implemented in R package *limma*.^41^ DE analysis was conducted to compare three conditions: bacterial infections versus controls, viral infections versus controls, and bacterial versus viral infections. Statistical significance was determined using a nominal p-value threshold of < 0.05, while biologically significant expression changes were identified based on log2-fold change (log2FC) thresholds of ≥ 1 or ≤ −1.

For the blood transcriptome data, differentially expressed genes (DEGs) were determined using an ensemble strategy combining two independent statistical frameworks: *limma* and *DESeq2*.^42^ To satisfy the underlying assumptions of each method, raw count data were handled separately: *DESeq2* was applied by fitting a negative binomial generalized linear model directly to the raw count data with normalization for sequencing depth, while *limma* was applied following a TMM normalization and log2 transformation to satisfy the assumptions of linear modelling. The results of two methods were combined by calculating the average log2FC value and the p-value according to the Fisher method^43^ as implemented in the R package *metap*.^44^ Statistical significance was determined using a Benjamini-Hochberg (BH) multiple testing corrected p-value threshold of < 0.05, while biologically meaningful expression changes were identified based on log2-fold change (log2FC) thresholds of ≥ 1 or ≤ −1. Significantly overrepresented pathways were identified via gene ontologies in DAVID.^45^

### Prioritization of potential diagnostic feature miRNAs

DE miRNAs in bacterial vs. viral infections were used to identify potential diagnostic features. Penalized sparce logistic regression as implemented in the R package *glmnet*^46^ was used to determine diagnostic signatures for bacterial infections, viral infections, and controls. A one-vs-all approach was used to build the model: the bacterial infection group served as the target group and was compared to the viral infection group and controls. The least absolute shrinkage and selection operator (LASSO) or L1 regularization was used to determine the smallest number of features while maintaining diagnostic accuracy. For optimal lambda coefficient calculation, leave-one-out cross-validation (LOOCV) was implemented. Accuracy, positive predictive value, the F1 score, and the Matthew correlation coefficient were calculated based on the confusion matrix to evaluate the performance of the model.^47^ ROC curve was generated via the R package *pROC*,^48^ along with the AUC score. To further refine the candidate biomarker pool, a univariate feature-screening phase was executed using an ANOVA F-test via the *SelectKBest*() in *scikit-learn*.^49^ The input features for this step were restricted to the miRNAs previously identified as altered between bacterial infection group and controls. To account for multiple comparisons during univariate screening, the raw p-values generated by the ANOVA F-test were adjusted using the Benjamini-Hochberg multiple testing correction method. miRNAs which achieved an adjusted p-value < 0.05 were considered statistically significant. Only features deemed important by both implemented methods were selected for further investigation. In addition to the one-vs-all approach, binary classification models were also constructed for pairwise comparisons between each infection group (bacterial vs. viral, bacterial vs. control, viral vs. control). A similar approach for feature selection as described in this chapter was applied to mRNA data as a complimentary analysis.

### miRNA:mRNA interaction network

To evaluate the potential interactions between miRNA biomarkers and differentially expressed genes in blood transcriptome, a miRNA target prediction analysis was performed. Three databases with distinct interaction detection methods were selected: one based on functional study results (*miRTarBase*),^50^ one using computational predictions (*DIANA-microT*),^51^ and one based on sequence similarity (*TargetScan*).^52^ mRNAs and miRNAs were considered to be linked if the interaction was identified in at least one of three databases. According to the widely accepted understanding, binding of miRNA to the corresponding mRNA induces translational inhibition and/or mRNA degradation.^53,54^ Therefore, miRNA interactions were only examined in down-regulated genes when comparing bacterial with viral infections. In addition to database analysis, correlation analysis was performed between the potential miRNA diagnostic feature and the genes predicted to interact with them. Pearson’s correlation coefficient was calculated using the R package *Hmisc*.^55^ The correlation was considered statistically significant if the coefficient was negative and with BH-adjusted p-value < 0.05. For visualizing the miRNA-gene interaction network, *Cytoscape* v3.10.2^56^ was used, while functional enrichment analysis was performed using the *ToppFun* v50 database (version from 30.11.2023)^57^ Results with a BH-adjusted p-value < 0.05 were considered statistically significant.

## Results

### Pilot study design

The cohort was recruited from pediatric patients from the Children’s Clinical University Hospital (Riga, Latvia) aged one month to 18 years with fever and infections confirmed by culture and clinical imaging for bacterial, or rapid antigen testing for viral infections. At the time of sample collection patients had no reports of known active urinary tract infections, pre-existing severe renal failure or acute kidney injury. Sample collection took place before the administration of antibiotic or antiviral therapies. The control group were children hospitalized for prior planned cardiac surgery. They had no reported fever or any other signs of infection and except for congenital heart defects such as VSD or ASD, had no other illnesses. Basic clinical information about the pilot study cohort is available in Table 1, while case-by-case diagnosis and information on infection severity is available in Supplementary Table S1. The outline of the main study workflow is described in Figure 1.

**Table 1.**
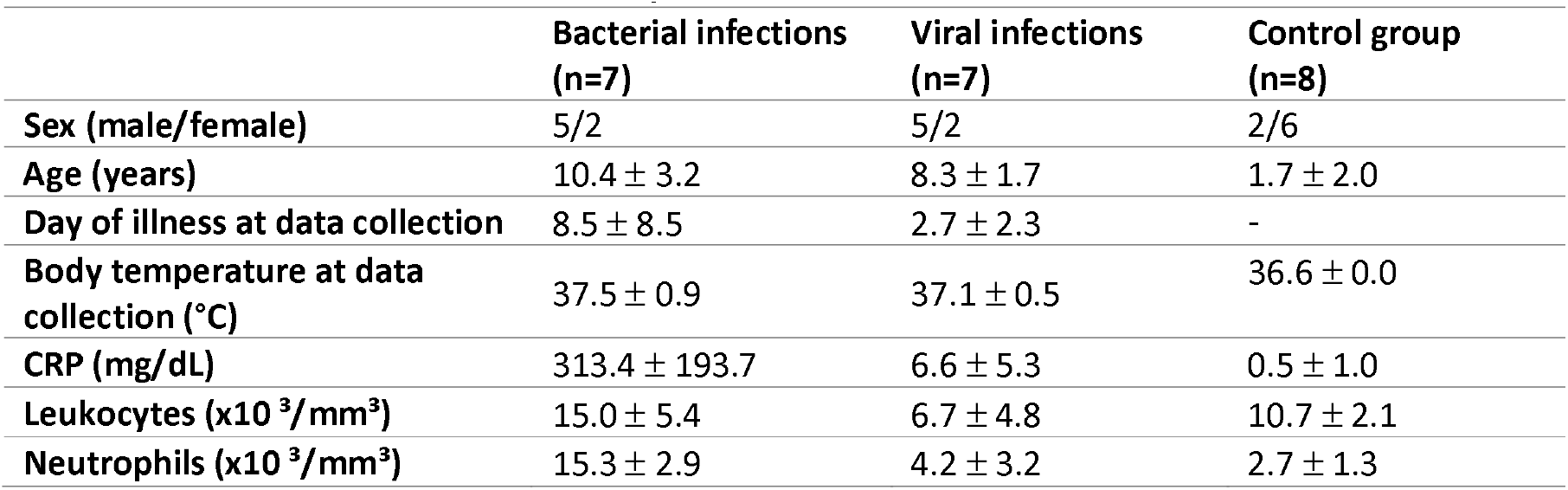
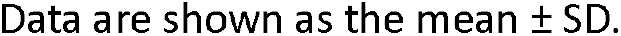
Basic clinical information on the study cohort.

|  | Bacterial infections<br>(n=7) | Viral infections<br>(n=7) | Control group<br>(n=8) |
| --- | --- | --- | --- |
| Sex (male/female) | 5/2 | 5/2 | 2/6 |
| Age (years) | 10.4 ± 3.2 | 8.3 ± 1.7 | 1.7 ± 2.0 |
| Day of illness at data collection | 8.5 ± 8.5 | 2.7 ± 2.3 | - |
| Body temperature at data collection (°C) | 37.5 ± 0.9 | 37.1 ± 0.5 | 36.6 ± 0.0 |
| CRP (mg/dL) | 313.4 ± 193.7 | 6.6 ± 5.3 | 0.5 ± 1.0 |
| Leukocytes (x10<sup>3</sup>/mm<sup>3</sup>) | 15.0 ± 5.4 | 6.7 ± 4.8 | 10.7 ± 2.1 |
| Neutrophils (x10<sup>3</sup>/mm<sup>3</sup>) | 15.3 ± 2.9 | 4.2 ± 3.2 | 2.7 ± 1.3 |

Data are shown as the mean ± SD.

**Figure 1.**
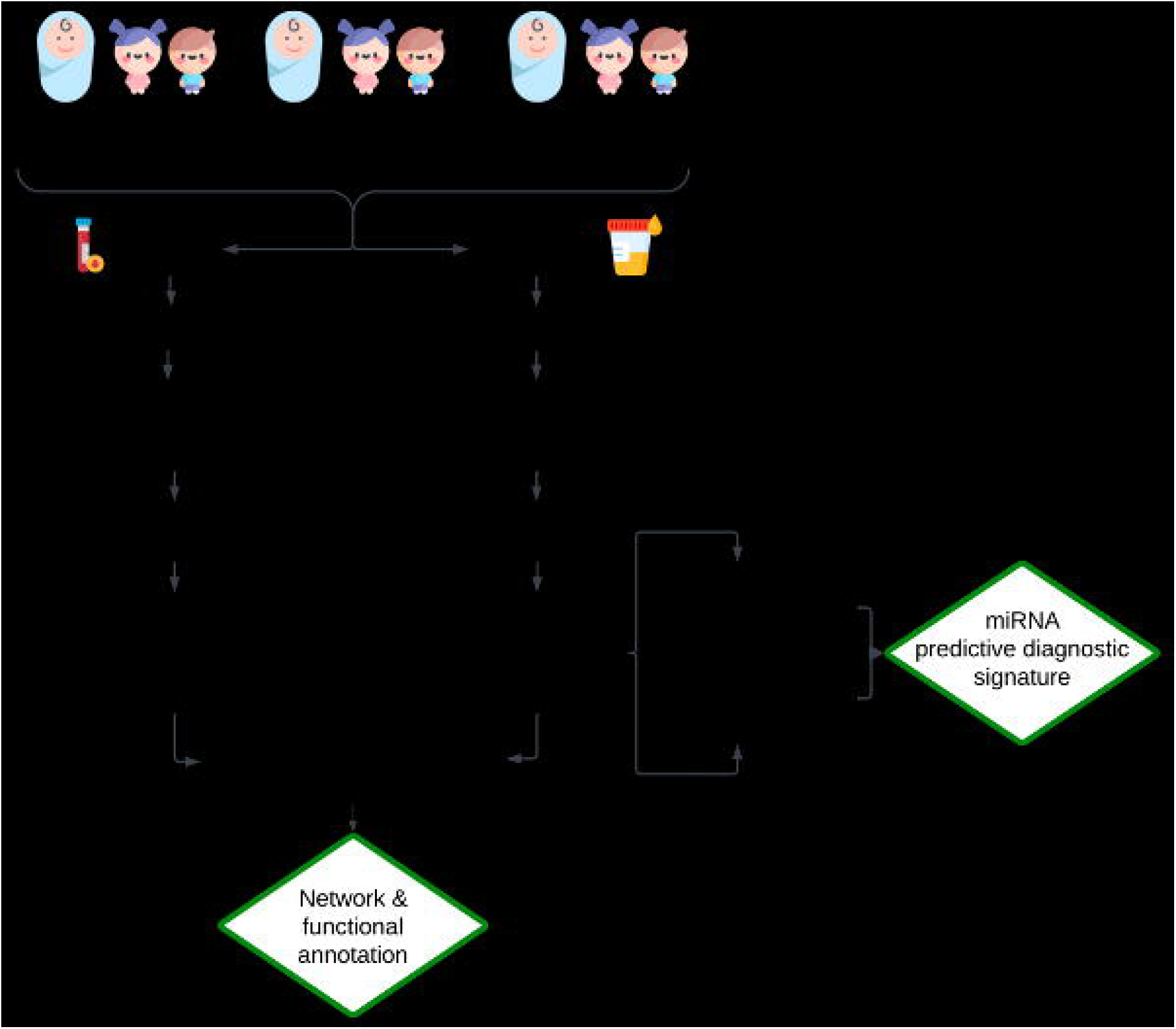
Outline of the main study workflow. 22 pediatric patients aged one month to 18 years were included in the study, comprising seven with proven bacterial infections, seven with proven viral infections, and eight controls without infection. RNA-seq was performed for both blood and urine samples, generating full transcriptome RNA-seq data and small RNA-seq data consecutively.

### miRNA differential expression profiles in bacterial, viral infections and controls

Differential expression analysis between the bacterial infection group (n = 7) and controls (n = 8) identified 64 nominally significant miRNAs (log2FC > 1 or < −1, nominal p < 0.05), with 44 (69%) up-regulated (median log2FC = 1.81, IQR = 1.42) and 20 (31%) down-regulated (median log2FC = −1.60, IQR = 0.85) in the bacterial group (Figure 2a). Similarly, between the viral infection group (n = 7) and controls (Figure 2b), 50 miRNAs were differentially expressed under these criteria, with 35 (70%) up-regulated (median log2FC = 1.64, IQR = 0.45) and 15 (30%) down-regulated (median log2FC = −1.58, IQR = 0.55). In the comparison of bacterial versus viral infections (Figure2c), 26 miRNAs were differentially expressed, evenly divided between up-regulated (13, 50%, median log2FC = 2.13, IQR = 0.87) and down-regulated (13, 50%, median log2FC = −1.33, IQR = 0.76). Rather than defining an established diagnostic signature at this stage, these nominally significant miRNAs were utilized as an exploratory baseline for diagnostic feature selection to identify potential multi-marker patterns to distinguish between bacterial and viral infections (see Supplementary Table S2).

**Figure 2.**
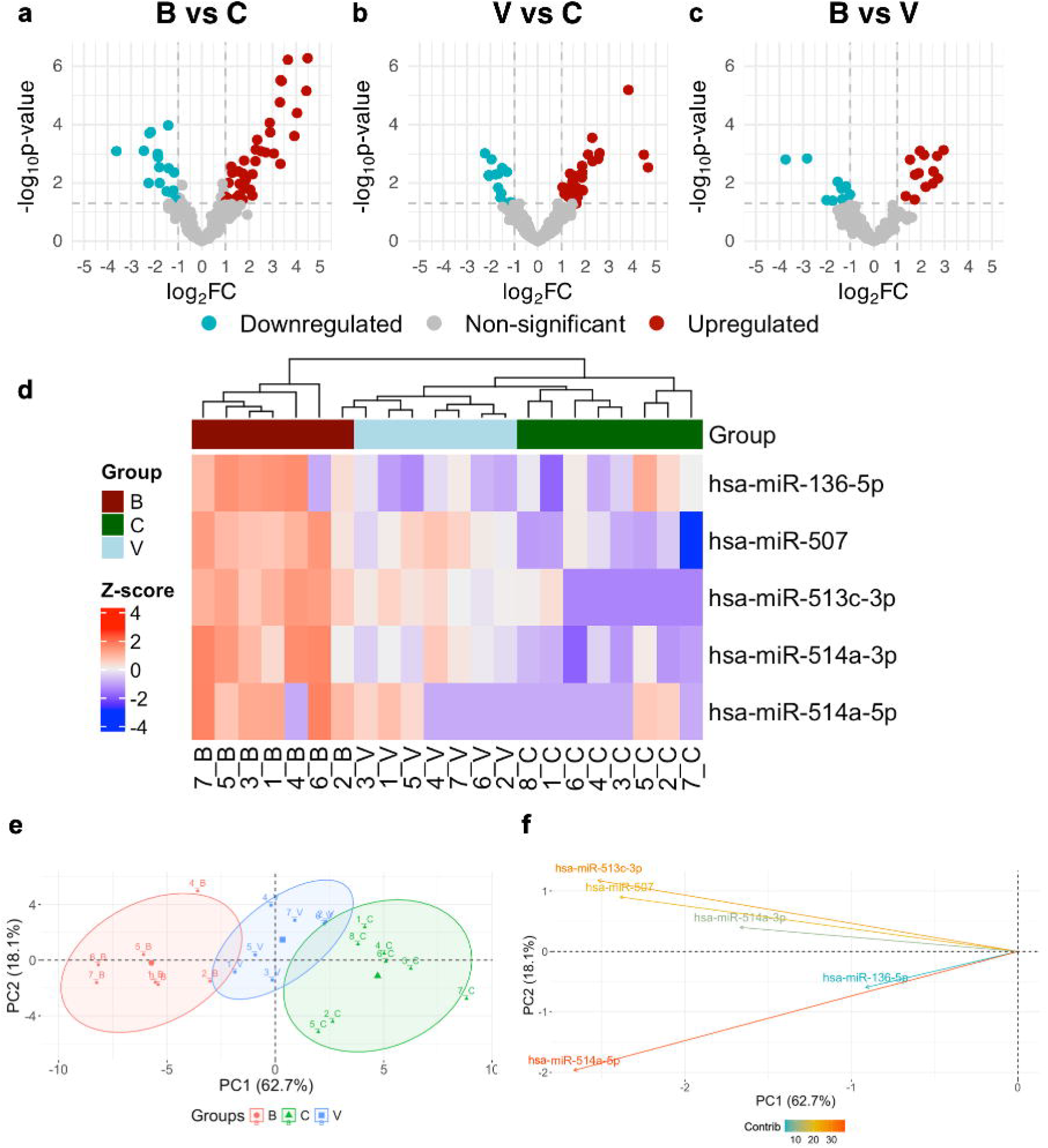
Exploring DE miRNAs and selecting urine miRNA signature for distinguishing infection etiology in febrile children. Volcano plots demonstrating identified differentially expressed (DE) miRNAs, comparing (A) bacterial infection group (n = 7) with controls (n = 8); (B) viral infection group (n = 7) with controls; (C) bacterial infection group against viral infection group. Up-regulated miRNA marked in red, down-regulated, in turquoise, miRNA not corresponding to the definition of DE miRNA in this study, in grey. (D) Heatmap along with agglomerative hierarchical cluster analysis using Z-score for selected miRNA biomarker candidates differentiating between bacterial and viral infections. Negative Z-score marked in blue, positive Z-score marked in red, Z-score reaching zero marked in white. In cluster analysis patients with bacterial infections marked in dark red, with viral infections – in light blue, controls – in green. (E) Principal component analysis with miRNA diagnostic signature selected based on LASSO penalized logistic regression and ANOVA analysis results. Samples from patients with bacterial infections (B) marked in red, from patients with viral infections (V) - blue, and from controls (C) - green. (F) Contribution of selected miRNAs to the principal components. The color scale represents the contribution to the first principal component in percents.

### Selection of miRNA signature to differentiate infection etiology

Two feature selection approaches were combined to select miRNA signatures that are distinct between groups: logistic regression model with LASSO regularization^58^ for bacterial versus viral infections and controls and variance analysis (ANOVA)^59^ between bacterial and viral infections.

LASSO regularized regression identified a 10-miRNA signature with non-zero coefficients (Supplementary Table S4), consisting of seven upregulated and three downregulated miRNAs in the bacterial infection group relative to the viral group.

The exploratory internal classification performance of this signature is presented in Table 2. The model achieved an apparent AUC score of 0.981 (Supplementary Figure S2), indicating high internal accuracy and strong discriminative performance across thresholds for this training set. However, given the small cohort size (N=22), the calculated 95% confidence intervals (CIs) reveal wide boundaries, particularly for the positive predictive value [0.421, 0.996]. This variance highlights the inherent instability of diagnostic estimates in small sample sizes, where the misclassification of a single sample can materially alter performance metrics. Consequently, these results must be interpreted as an internal exploratory performance rather than a validated diagnostic output.

**Table 2.**
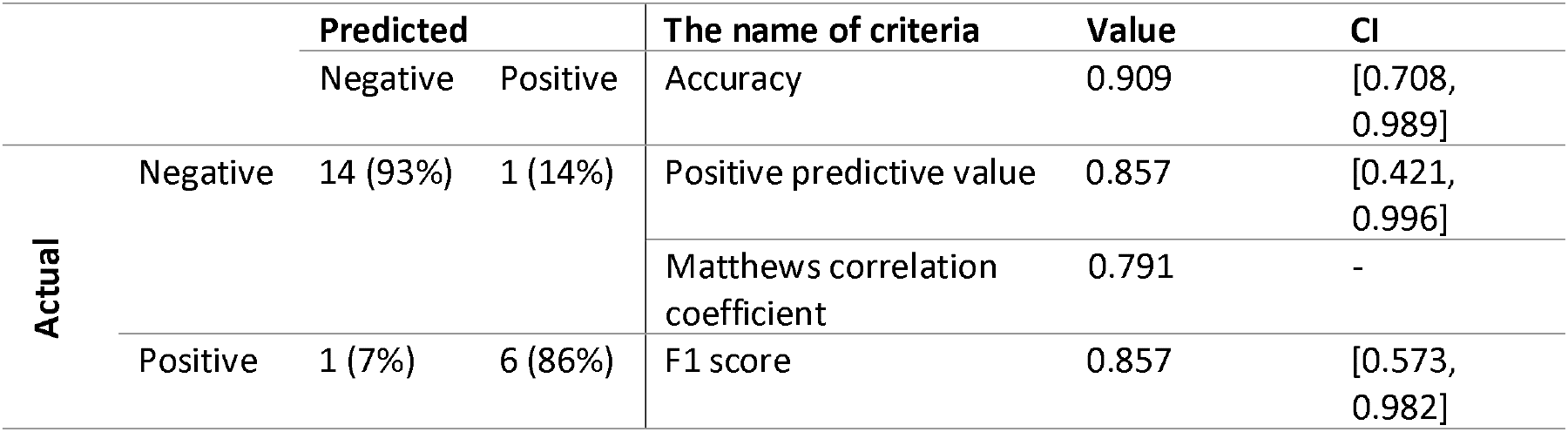
Internal classification performance and confusion matrix of the LASSO regression model.

As a result of the additional application of ANOVA for diagnostic feature evaluation, 11 miRNAs were identified as significant (BH-adjusted p-value < 0.05), as shown in the Supplementary Table S4. Nine of these miRNAs are up-regulated, while two are down-regulated.

Combining both methods described above, six miRNAs identified as potential diagnostic features were selected: *hsa-miR-136-5p, hsa-miR-513c-3p, hsa-miR-514a-5p, hsa-miR-514a-3p, hsa-miR-1-3p, hsa-miR-507. hsa-miR-1-3p* was excluded from downstream analysis due to its low expression level compared to other selected miRNAs (10,3 ± 5,5 mean relative expression). The overlook on the selected miRNA signature is available in Table 3.

**Table 3.** Overview of selected miRNA signature for distinguishing between bacterial and viral infections.

| Name of miRNA | Up- or down-regulated (↑/↓) | Relative expression level |
| --- | --- | --- |
| *hsa-miR-136-5p* | ↑ | $100.1 \pm 19.0$ |
| *hsa-miR-507* | ↑ | $243.6 \pm 40.0$ |
| *hsa-miR-513c-3p* | ↑ | $19.5 \pm 13.0$ |
| *hsa-miR-514a-3p* | ↑ | $870.5 \pm 146.5$ |
| *hsa-miR-514a-5p* | ↑ | $45.8 \pm 16.0$ |

Overview on miRNA signature selected via LASSO regularization (based on LASSO coefficient) and ANOVA (based on p-value), along with summary on expression levels and up-or down-regulation.

Hierarchical cluster analysis (see Figure 2d) was performed to visualize the expression profiles of the selected miRNA signature across the patient cohort. The resulting heatmap illustrates distinct expression patterns between the groups, reflecting biological variance between the bacterial infection group, the viral infection group, and controls. The results of the hierarchical agglomerative clustering (top of Figure2d) show greater similarity between the viral infection group and the control group than between the bacterial infection group and the other comparison groups. Notably, a sample from the bacterial infection group (Sample 2) is clustered with the patients with viral infection, which is also observed in Figure 2e in the principal components analysis.

Figures 2e and 2e show the results of the principal component analysis (PCA), using only the selected miRNA diagnostic signature. The first principal component explains 62.7% of the data variability and the second component explains 18.1%. First two principal components are sufficient to explain at least 80% of the data variability, while using three components allows explaining 91.4% of the variability. Figure 2f shows the contribution of each individual miRNA to the principal components. The largest contributions to the first principal component come from *hsa-miR-514a-5p* (31.2%), *hsa-miR-513c-3p* (27.9%), and *hsa-miR-507* (25.0%), while for the second component, *hsa-miR-514a-5p* represents 58.8%. It can be concluded that *hsa-miR-514a-5p* provides the largest contribution to explaining the variance captured by the selected signature. Because these visualizations are based on features pre-selected from the same dataset, the clear cluster separation is descriptive of the signature’s underlying variance rather than independent validation of classifier performance.

To provide broader context, a supplementary binary evaluation was performed using separate LASSO regression models to classify candidate profiles across individual cohort pairs (Supplementary Figure S1; Supplementary Table S3). Direct classification between bacterial and viral groups yielded an apparent internal accuracy of 0.929 (95% CI: 0.661–0.998) and an F1 score of 0.923 (95% CI: 0.621–0.996). When comparing the bacterial or viral cohorts individually against healthy controls, both models demonstrated identical exploratory metrics, achieving an apparent internal accuracy of 0.867 (95% CI: 0.595–0.983) and an F1 score of 0.875 (95% CI: 0.584–0.975). Due to unnested feature screening, these metrics serve purely as exploratory, internal performance descriptors within the supplementary material.

### Full transcriptome differential expression profiles and functional analysis in bacterial, viral infections and controls

Differential expression (DE) analysis between the bacterial patient group and controls (Figure 3a) identified 5,860 differentially expressed genes (DEGs), with 2,811 (47.97%) up-regulated and 3049 (52.03%) down-regulated. The up-regulated genes had a median log2FC of 1.50 (IQR = 0.81), while the down-regulated genes had a median log2FC of −1.38 (IQR = 0.54). Similarly, DE analysis between the viral patient group and controls (Figure 3b) revealed 2,218 DEGs, with 1,050 (47.34%) up-regulated and 1,168 (52.66%) down-regulated. The median log2FC for the up-regulated genes was 1.42 (IQR = 0.64), and for the down-regulated genes, it was −1.48 (IQR = 0.63). In the comparison between the groups of bacterial and viral patients (Figure 3c), 3,675 DEGs were identified, with 2,213 (60.22%) up-regulated and 1,462 (39.78%) down-regulated. The median log2FC for the up-regulated genes was 1.42 (IQR = 0.65), while for the down-regulated genes it was −1.37 (IQR = 0.57).

**Figure 3.**
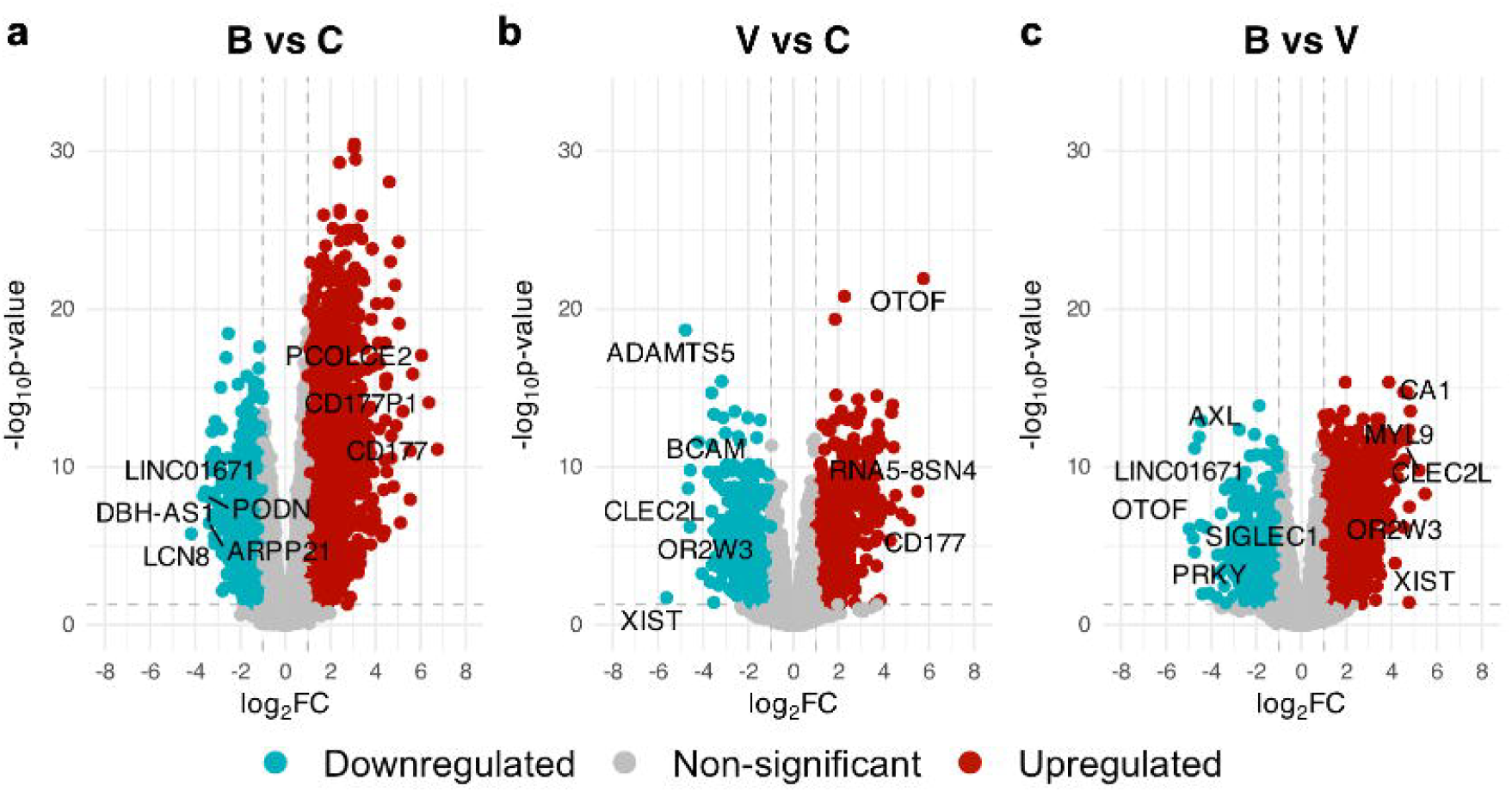
Volcano plots demonstrating identified differentially expressed (DE) transcripts in blood. (A) Comparing the bacterial infection group (n=7) against controls (n=8); (B) viral infection group (n=7) against controls (B); (C) bacterial infection group against the viral infection group. Up-regulated transcripts marked in red, down-regulated – in turquoise, mRNA not corresponding to the DE mRNA definition of this study – in grey. The top five up-regulated and down-regulated mRNA names are marked next to the corresponding plot points.

Using the identified DEGs, functional analysis was performed using gene ontologies (GO) in DAVID.^45^ All the observed pathways (overview provided in Table 4) are not only concordant with the known biology of infection, but also highly significant in the used dataset, such as adaptive immune response, inflammatory response, and defense response to virus in case of viral infections.

**Table 4.**
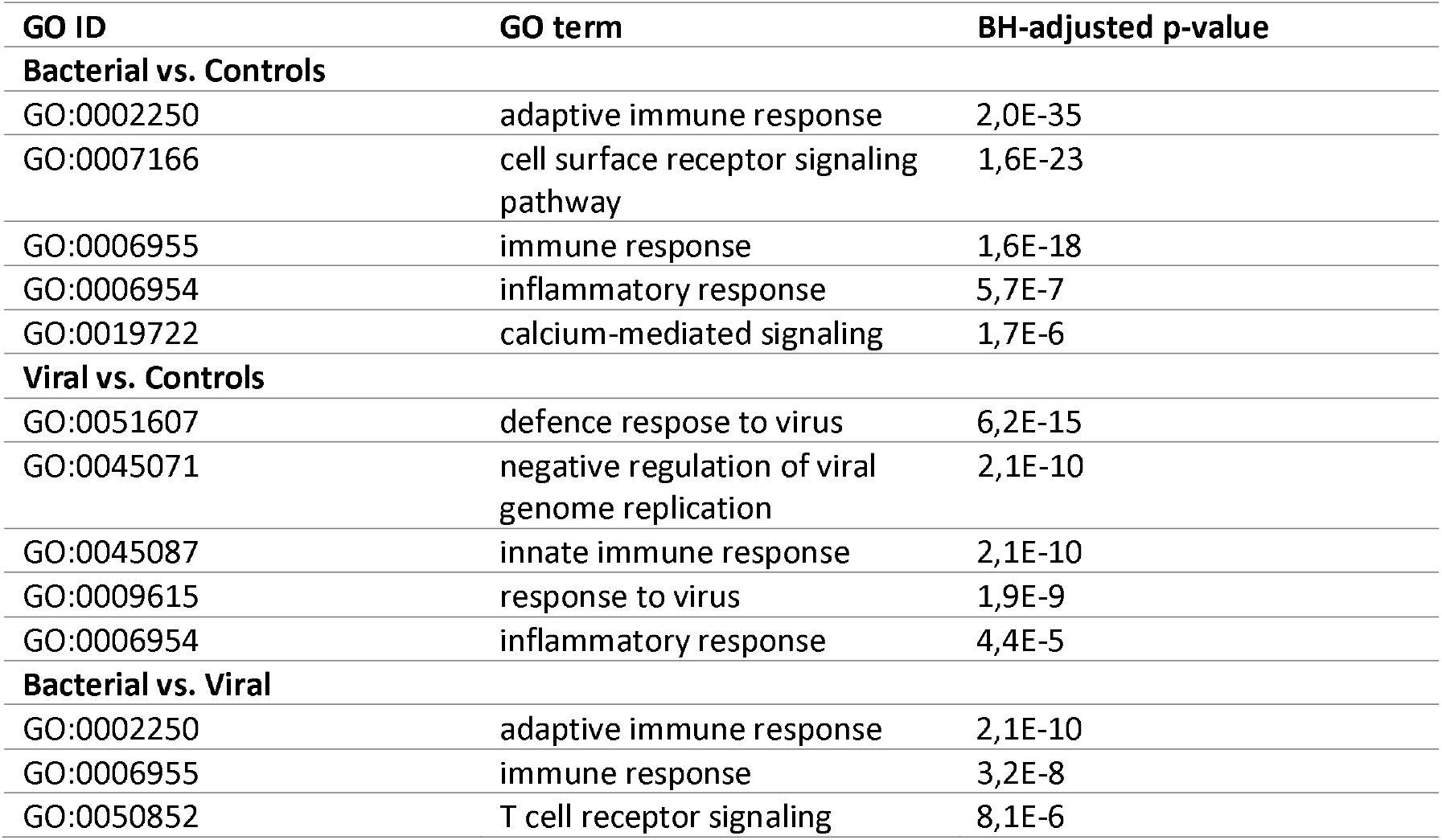
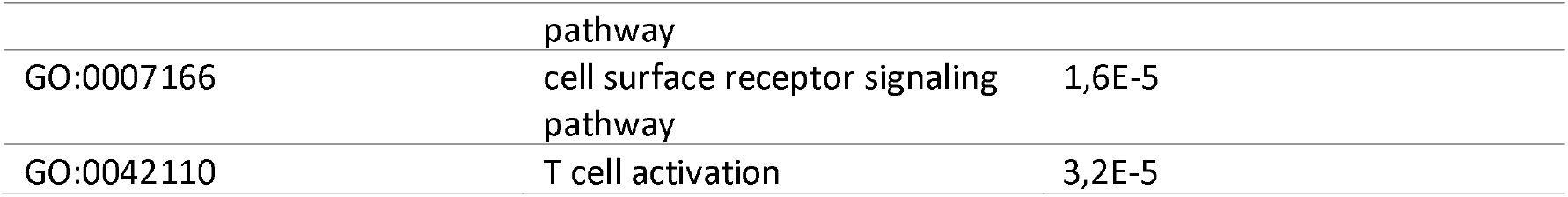
Top 5 upregulated GO terms associated with DEGs in three comparison groups derived from upregulated genes.

| GO ID | GO term | BH-adjusted p-value |
| --- | --- | --- |
| **Bacterial vs. Controls** |  |  |
| GO:0002250 | adaptive immune response | 2,0E-35 |
| GO:0007166 | cell surface receptor signaling pathway | 1,6E-23 |
| GO:0006955 | immune response | 1,6E-18 |
| GO:0006954 | inflammatory response | 5,7E-7 |
| GO:0019722 | calcium-mediated signaling | 1,7E-6 |
| **Viral vs. Controls** |  |  |
| GO:0051607 | defence response to virus | 6,2E-15 |
| GO:0045071 | negative regulation of viral genome replication | 2,1E-10 |
| GO:0045087 | innate immune response | 2,1E-10 |
| GO:0009615 | response to virus | 1,9E-9 |
| GO:0006954 | inflammatory response | 4,4E-5 |
| **Bacterial vs. Viral** |  |  |
| GO:0002250 | adaptive immune response | 2,1E-10 |
| GO:0006955 | immune response | 3,2E-8 |
| GO:0050852 | T cell receptor signaling | 8,1E-6 |

|  | pathway |  |
| --- | --- | --- |
| GO:0007166 | cell surface receptor signaling | 1,6E-5 |
|  | pathway |  |
| GO:0042110 | T cell activation | 3,2E-5 |

To ensure that the findings of this study align with previous research, differentially expressed genes (DEGs) for the bacterial vs. control and viral vs. control comparison groups were evaluated against published mRNA diagnostic signatures for bacterial and viral infections.^11,60–66^ This literature review identified 121 matched genes of the 245 reported in signatures listed for bacterial infections and 63 matched genes of the 276 reported in signatures for viral infections.

In addition to exploring the urinary miRNA diagnostic signature for bacterial infections versus viral infections and controls, a similar feature selection approach described in Methods section 2.5 was applied to blood mRNA data. This blood transcriptomic analysis was conducted purely as a contextual effort to complement the urinary data, rather than as a parallel diagnostic signature-development framework. The results of this exploratory analysis are listed in Supplementary Figure S3 and Supplementary Table S5. Consistent with its exploratory nature, the derived blood mRNA signature for bacterial versus viral infections did not demonstrate sufficient ability to differentiate the infection groups (results not shown). For the comparison of bacterial infections versus controls, a four-gene signature was derived (*FCRL3, WNT10B, PODN*, and *MMP23B*), while for the viral infections versus controls group, a single gene (*ADAMTS5*) was shown to differentiate between patients and the control cohort.

### miRNA:mRNA regulatory network of signature miRNA and differentially expressed genes

To explore the broader systemic landscape during the infection process, the relationship between the candidate urinary miRNA signature and the blood DEGs of the same patients was contextually mapped utilizing a theoretical target prediction analysis to screen for parallel pathways across compartments. Since miRNA conducts gene regulation by mediating translation inhibition^15^ through the promotion of translational inhibition and mRNA degradation, for correlation analysis only down-regulated genes were included. 34 pairs of miRNAs and DEG signature with statistically significant (BH-adjusted p-value < 0.05) negative correlation were discovered using Pearson’s correlation.

Within the selected 5-miRNA signature, three miRNAs showed negative correlations, mapping to potential parallel molecular pathways within the blood DEG dataset: *hsa-miR-507* (25 pairs), *hsa-miR-514a-3p* (3 pairs), and *hsa-miR-513c-3p* (6 pairs). The full list of these putative interactions is available in Supplementary Table S6. The cross-compartment miRNA:mRNA network was visualized using these correlated pairs alongside functional annotations from the ToppFun database to outline parallel biological processes (see Figure 4). Characterized pathways focusing on innate and adaptive immune response framework development were highlighted within the network, specifically capturing lymphocyte differentiation, cytokine-cytokine receptor interactions, and the regulation of T-cell activation.

**Figure 4.**
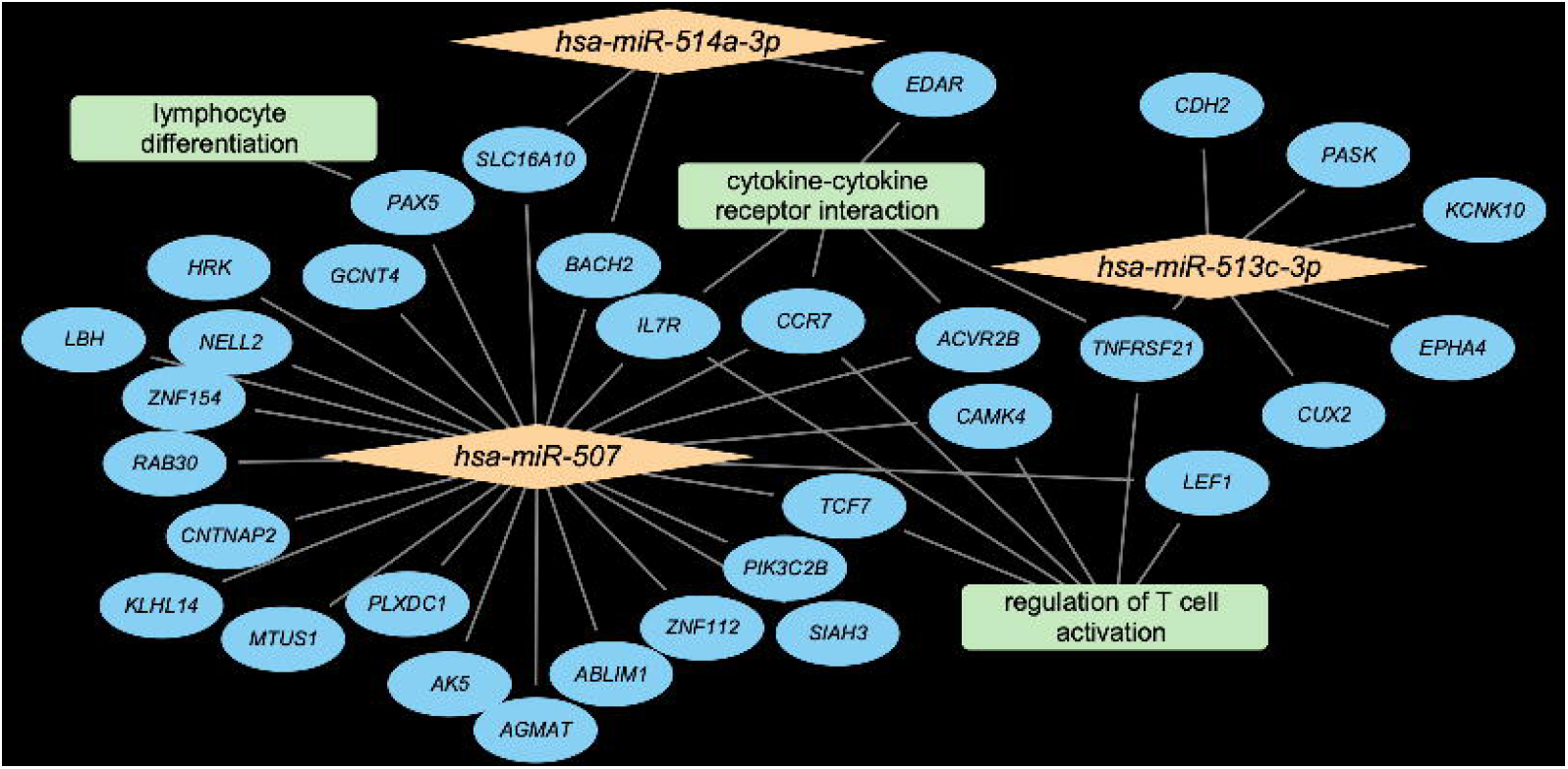
Integrated analysis and interaction network of signature miRNAs in urine and respective blood mRNA. miRNAs shown in orange, genes in blue, gene functions in green. The network was visualized using *Cytoscape* v3.10.2.^56^

Furthermore, functional enrichment analysis was performed for 21 miRNAs that were differentially expressed in bacterial or viral infection patient groups compared to controls; however, they were not DE when comparing patient groups (the full list is available in Supplementary Table S7). The main goal of this analysis was to accompany the information gained after creating the interaction network in Figure 4 on the role of miRNA in the regulation of the immune response and to discover which miRNA-regulated pathways may not be influenced by the etiology of the infection. Four of the miRNA mentioned *(hsa-miR-204-3p, hsa-miR-27b-5p, hsa-miR-508-3p, and hsa-miR-508-5p*) are part of non-canonical NF-κB signal pathway, as well as the regulation of the synthesis of IL-1 along with *hsa-miR-708-3p. hsa-miR-204-3p, hsa-miR-27b-5p, hsa-miR-708-3p, hsa-let-7f-1-3p and hsa-miR-200c-3p* (the only miRNA down-regulated out of all mentioned above) are a part of cytokine synthesis regulation. All the mentioned pathways are part of the innate immune response and initiate and sustain the inflammatory process regardless of the etiology of the infection.^67–69^

## Discussion

The potential applications and importance of urine biomarkers in pediatric care is a highly promising area of research. Urine can be collected easily and non-invasively.^70^ Considering the lack of reliable and rapid infection etiology diagnostic methods, the search for new biomarkers is an important step to improve the quality of healthcare.^71^ The use of transcriptome and specifically miRNAs in diagnostics has considerable potential as a tool to leverage in personalized medicine approaches. High stability of miRNA in biological fluids suggests its high biomarker potential.^72^ Furthermore, the role of miRNAs as upstream regulators indicates that a single miRNA biomarker may be sufficient to detect systemic changes such as activation and deactivation of immune pathways, making miRNAs even more attractive candidates as disease diagnostics tools.^17^ The studies of infection biomarkers up to date have primarily focused on blood mRNA transcripts, but growing recognition of miRNAs as prominent regulators of various biological pathways and diseases made us turn our attention to miRNAs.

The DE analysis performed highlighted significant differences between the expression profiles of patient groups and controls. It suggests not only the importance of miRNAs in the regulation of the immune response and its involvement in the infection process,^15,73,74^ but also the fact that these complex changes could potentially be detected in urine which has not previously been reported. When comparing DE miRNA in the viral infection group against controls and the bacterial infection group against controls, it was found that multiple up-regulated and down-regulated miRNAs overlap between these. It may suggest the role of DE miRNAs in the regulation of nonspecific immune response that is not related to the etiology of infection. The latter is partially confirmed by comparison of miRNAs that are DE between patient groups and controls, however, are not DE when comparing between patient groups: all the relevant pathways discovered are related to innate immune response and inflammation initiation regardless of infection etiology. It has been shown that fever in rat models activates additional inflammation inducing pathways that may be miRNA-regulated^75^ which may suggest that a similar process is happening in a human host organism. Because this pilot study relied on a nominal p < 0.05 significance cutoff for DE analysis without strict correction for multiple comparisons, these captured adjustments serve strictly as a relaxed filtering threshold. This approach was implemented to maximize downstream data retention, focusing purely on identifying potential multi-marker signature patterns for future, larger-scale validation rather than defining a definitive, established diagnostic panel.

During the feature selection process for diagnostic signature that could distinguish between bacterial and viral infections, five up-regulated miRNA in bacterial versus viral infections patient groups (specifically *hsa-miR-136-5p, hsa-miR-513c-3p, hsa-miR-514a-5p, hsa-miR-514a-3p, and hsa-miR-507*) were selected as candidates of a potential diagnostic signature to differentiate the etiology of infection. Two feature selection methods were combined: logistic regression with LASSO regularization^58^ and ANOVA.^76^ The rationale behind this combination was the widespread use of LASSO regularized logistic regression in multi-omics data analysis and the comparative simplicity of ANOVA and its ability to reduce the occurrence of Type I and Type II errors in feature selection, particularly in small datasets with a large number of features.^59^ The implemented logistic regression was using the one-vs-all principle, which is widely used in the selection of clinically significant feature: even when multiple groups are present, the samples are divided into the “relevant” class (which detection is the most significant for clinical interpretation) and the “other” class.^47^ The prioritized candidate miRNA signature demonstrated encouraging apparent internal performance in distinguishing bacterial from non-bacterial profiles within this exploratory cohort.

Although the focus of this study is the exploratory discovery of the potential diagnostic signature of miRNA for bacterial and viral infections, blood derived mRNAs from the same patients were used to compare the discovered DEG profile with DEGs reported in various studies, and report on significantly overrepresented pathways. Most pathways discovered via functional annotation are connected to immune response (adaptive immune response, inflammatory response, defense response to virus, etc.) which corresponds with previous findings of the above-mentioned studies.^11,60–66^ However, when comparing the DEG profile observed in this study with other studies, it has been concluded that only 49% and 23% of genes overlap between bacterial infections vs. controls and viral infections vs. controls accordingly. This highlights the fact previously described in the systematic comparison of published gene signatures for bacterial and viral infections: the gene signatures will vastly vary depending on the validation population and will perform best in the studied demographic specifically.^14^ The genes highlighted in this study have reports on playing a role in immune system functions, namely *WNT10B* and *FCRL3*.^77,78^ *ADAMTS5* has been mentioned as a virus-specific immunity regulator.^79^

Potential parallel pathways between candidate urinary miRNAs and blood DEGs within the same patient cohort were computationally evaluated across these two distinct biological fluids: urine and blood. Most of the literature on infection transcriptomics focuses on the whole blood transcriptome, and the additional objective of this exploratory pilot study was to investigate the whole blood transcriptome given their prognostic value highlighted in other publications.^70,72,80^ As there have been studies that have found the connection between the blood and urine transcriptome and miRNA content, which can be both cell-free and part of exosome cargo, our aim was to investigate if complex immune changes during the infection could also be potentially detectable in urine, as RNA in urine could be derived from RNA in bloodstream that has been filtered by the glomerulus.^81^ Although there has been no direct proof of this statement to date, it has been observed that urine may contain tumor-derived nucleic acids from the non-urinary tract^82^ and fetal DNA.^83^ The cross-compartment correlations and target-prediction database results presented here serve as an exploratory, hypothesis-generating tool rather than establishing direct physical regulatory interactions between urinary miRNAs and circulating transcripts. The results of this study further develop the possible connection between the blood and urine transcriptome and miRNA content, highlighting parallel biological axes triggered by the host response.

The search of the literature to compare findings related to miRNA DE profiles in bacterial and viral infections yielded limited results, mainly due to the scarcity of relevant studies. Poore et al. (2018) study on miRNA diagnostic signatures for bacterial and viral infections in adult blood has not discovered a single statistically significant (FDR ≤ 0.01) DE miRNA between patients with viral infections and controls. When applying the same threshold to the results of this study, one miRNA is selected, *hsa-miR-508-3p*, which is upregulated in this comparison group. According to the literature, it is involved in apoptosis regulation which can be a part of non-specific immune response;^84^ however, there is no further information on its connection to infectious diseases. When comparing DE miRNA lists of bacterial infections vs. controls and bacterial vs. viral infections, nine and two of DE miRNAs from this study match with the results of Poore et al.

Until now, there have been limited reports on the regulatory role of the aforementioned diagnostic feature miRNA. Expression of *hsa-miR-136-5p* mediated the synthesis of molecules involved in inflammation formation - IL-1β, IL-6, TNF-α, IFN-α, IKKβ and NF-κB - in rat models.^85^ All the mentioned compounds are involved in the formation of an innate immune response. *hsa-miR-507* has been associated with chronic kidney disease, which may indicate its role in the inflammation response in the urinary tract.^22^ *hsa-miR-513c-3p* is mentioned as a candidate biomarker for the distinction between HIV-1 and HIV-2 viral infections.^86^ hsa-miR-514a-3p is involved in autophagy, which is an important part of the anti-inflammatory response.^87^

As mentioned previously, the literature on miRNA infection markers is limited, restricting the possibilities of comparing results with other papers or validating the mentioned finds in independent cohorts. The study by Poore et al. (2018) mentioned above highlights five miRNAs to distinguish bacterial and viral respiratory infections (pneumonia caused by *Streptococcus pneumoniae* infection and H3N2 flu) - *hsa-miR-942-5p, hsa-miR-342-5p, hsa-miR-503-5p, hsa-miR-199-5p* and *hsa-miR-30a-5p*. Of five, none of the described miRNAs were selected as an important diagnostic signature. Logistic regression highlighted *hsa-miR-30a-3p*, which comes from the 3p arm of the same pre-miRNA as *hsa-miR-30a-5p* and may possess similar functions.^88^ The observed differences between the study results could be explained by the fact that the overall quantity of miRNA in urine is much lower than that in blood,^89^ which may explain the different profile of discovered miRNAs. In addition, it is not known how the miRNA expression in blood and urine correlates, so it is possible that diagnostic signatures discovered by Poore and colleagues do not translate into expression in urine in the same way.

While the linear regression method used during this study showed good internal performance in distinguishing bacterial infections from viral infections and controls, the same approach did not achieve the same results for determining viral infections from bacterial infections and controls. This corresponds to results from previously cited findings by Poore et al. (2018). The study by Hu et al. (2013), in addition, state that they have not found significant differences between gene expression profiles in afebrile virus-infected and afebrile non-infected children, which may prove that it is much harder to detect gene expression changes in virus-infected pediatric cohorts than in bacteria-infected cohorts.

While the direct differentiation of bacterial and viral etiologies was the primary objective of this pilot study, expanding the analysis to alternative binary classification tasks - such as distinguishing infectious cohort from healthy controls - provides critical context for future clinical deployment frameworks. To address these distinct classification questions within our cohort, separate exploratory LASSO logistic regression models were evaluated to assess candidate signatures across individual pairs, including bacterial versus control and viral versus control profiles. While these models demonstrated encouraging internal exploratory capabilities—yielding apparent accuracies of 0.867 for control comparisons and 0.929 for the direct bacterial versus viral classification—the lack of an independent non-infectious febrile control group prevents a true evaluation of a generalized ‘infection versus no infection’ baseline. Consequently, completely disentangling etiology-specific markers from shared, non-specific inflammatory pathways will require larger-scale validation studies that incorporate pathologically matched, non-infectious febrile control cohorts.

We acknowledge that our study, while showing promising results, has some limitations. This pilot study utilizes a small cohort which may lead to the overfitting of the model. While LOOCV was used to evaluate this, the preliminary differential expression analysis and initial feature selections were performed on the full dataset prior to the cross-validation loop. This lack of fully nested feature selection introduces a risk of data leakage, meaning our reported metrics should be interpreted as an apparent, internal exploratory performance.

Additionally, samples were collected at a single time point, which varied depending on the patient’s hospital admission day. A more standardized collection protocol, involving multiple time points during the infection, could reflect greater variability and enhance the model’s scaling to other cohorts. While patients with different bacterial infections were included to improve generalization, this diversity may have limited the identification of specific miRNAs. On the contrary, the viral infection cohort included only patients with influenza, which could have the opposite effect. A larger and more diverse validation cohort would ensure the robustness of the identified diagnostic features and their suitability for clinical application. Additionally, the relationship between urine and blood transcriptomes remains unexplored, making it challenging to determine the scalability of the connections observed in this study.

Furthermore, baseline imbalances in clinical characteristics between the cohorts, specifically regarding patient age and the day of illness prior to sampling, introduce potential sources of unexpected biological variability. In this pilot study, variations in the duration of illness were partially driven by specific clinical outliers, such as a patient who developed a secondary infection during a prolonged intensive care unit stay. Because this study is underpowered to perform robust multivariable covariate adjustments or matching protocols, these imbalances remain unadjusted. Future large-scale verification studies must implement strict age-matching strategies and standardized symptom duration enrolment to minimize confounding host-response variations.

To the best of our knowledge, this is the first study to use urine miRNA for the prioritization of the diagnostic signature for infections that are not related to the urinary tract. The role of miRNA in the regulation of immune response is not well studied^74^ therefore, it requires further investigation. The candidate urinary miRNA signatures identified in this exploratory study provide a potential foundation for future molecular diagnostic research, serving as a non-invasive tool to investigate host-response mechanisms across distinct infectious etiologies. In emergency care, a quick and accurate differentiation between bacterial and viral infections is crucial for making decisions about the appropriate treatment protocols. The findings on miRNA expression in urine suggest the possibility of using urine to characterize systemic processes not only in the context of metabolomics,^91^ but also in transcriptomics. To validate these preliminary insights, future studies should evaluate a larger patient cohort and directly compare this method against existing diagnostic techniques.

## List of abbreviations

ANOVA: Analysis of variance
BH: Benjamini-Hochberg multiple testing correction
bp: Base pair
CI: Confidence interval
CRP: C-reactive protein
DAVID: Database for Annotation, Visualization, and Integrated Discovery
DE: Differential expression
DEG: Differentially expressed gene
FC: Fold change
FDR: False discovery rate
FN: False negative
FP: False positive
GO: Gene ontology
HIV: Human immunodeficiency virus
IFN: Interferon
IKKβ: Inhibitor of nuclear factor kappa-B kinase subunit beta
IL: Interleukin
IQR: Interquartile range
LASSO: Least absolute shrinkage and selection operator
LOOCV: Leave-one-out cross-validation
AUC: Area under curve
miRNA: microRNA
mRNA: messenger RNA
NF-κB: Nuclear factor kappa-B
nt: Nucleotide
PCA: Principal component analysis
PPV: Positive predictive value
RNA-seq: RNA sequencing
TLR: Toll-like receptors
TMM: Trimmed mean of the log expression ratios (M values)
TN: True negative
TNF-α: Tumor necrosis factor α
TP: True positive
ROC: Receiver operating characteristic curve

## Declarations

### Ethics approval and consent to participate

The study was approved by the Ethics Committee of Riga Stradins University (Nr.6-2/4/ 2, dated 25.04.2019) and was carried out in accordance with the Declaration of Helsinki. All legal guardians of the recruited subjects gave their written informed consent to participate.

### Consent for publication

Not applicable. The manuscript does not contain any individual personal data.

### Availability of data and materials

The gene expression datasets used and analyzed during the current study is available in Gene Expression Omnibus under accession numbers GSE290693 and GSE290432 as well as in Riga Stradins University Dataverse from https://doi.org/10.48510/FK2/GZBT9C. The code used to generate and analyze the data is available on GitHub, DOI: <u>10.5281/zenodo.14845445</u>

### Authors’ contributions

DC and GZN contributed to the conception, design and securing the funding; GZN contributed to the collection of study materials and clinical data. TK and BV contributed to data bioinformatical pre-processing, analysis, interpretation and visualization. TK drafted the first version of the manuscript; all authors contributed to manuscript editing. All the authors have read and approved the final version of the manuscript.

## Acknowledgements

We would like to thank the nurses, patients and their legal guardians for participating in this study. We would also personally like to thank our colleague Līvija Bārdiņa for technical support in preparation for the data analysis.

