## Supplementary Information for "Exploratory Study on Blood and Urine Transcriptomes for Identifying miRNA Biomarkers in Pediatric Febrile Bacterial and Viral Infections"

**Contents:**

Supplementary Table 1. Clinical information on bacterial and viral group patients used in the study (n = 14).

Supplementary Table 2. A list of differentially expressed miRNAs comparing bacterial to viral infections.

Supplementary Table 3. Confusion matrix and associated internal performance metrics for models for the binary classification of urine miRNA between comparison groups.

Supplementary Table 4. Overview of selected miRNA potential candidate diagnostic signatures.

Supplementary Table 5. Internal classification performance and confusion matrix of LASSO regularized regression for the binary classification of blood mRNA between the bacterial infections group and controls.

Supplementary Table 6. The results of the Pearson's correlation analysis for miRNA and their respective regulated genes.

Supplementary Table 7. A list of miRNAs that are differentially expressed in the bacterial or viral patient group compared to the controls, but not differentially expressed when comparing the patient groups to each other.

Supplementary Figure 1. Selection of miRNA binary signatures from urine to distinguish infection etiology in febrile children.

Supplementary Figure 2. ROC curve for model used for miRNA selection.

Supplementary Figure 3. Selection of mRNA blood-based binary signatures to distinguish infection etiology in febrile children.

**Supplementary Table S1.** Clinical information on bacterial and viral group patients used in the study (n = 14).

| Patient ID | Bacterial | Viral | ICU | Respiratory support | Inotrope requirement | Length of stay, days | PELOD |
| --- | --- | --- | --- | --- | --- | --- | --- |
| 1_B | Burn wound infection with MSSA | - | yes | no | no | 18 | 0 |
| 2_B | *Staphylococcus aureus* from pleural fluid culture; pneumonia with empyema on chest x-ray | - | yes | yes | no | 17 | 4 |
| 3_B | *Streptococcus pyogenis* wound infection with sepsis | - | yes | no | yes | 6 | 2 |
| 4_B | *Nesseria meningitidis* sepsis and meningitis | - | yes | yes | no | 10 | 3 |
| 5_B | Sepsis, pneumonia on chest x-ray | - | yes | yes | yes | 12 | 9 |
| 1_V | - | Influenza type B, positive antigen test | no | no | no | 2 | 0 |
| 2_V | - | Influenza type B, positive antigen test | yes | no | no | 2 | 9 |
| 7_V | - | Influenza type B, positive antigen test | no | no | no | 7 | 0 |
| 3_V | - | Influenza type B, positive antigen test | yes | yes | no | 7 | 8 |
| 4_V | - | Influenza type B, positive antigen test | no | no | no | 2 | 0 |
| 6_B | Pneumonia with empyema | - | yes | yes | no | 28 | 2 |
| 5_V | - | Influenza type A, positive antigen test | no | no | no | 2 | 0 |
| 6_V | - | Influenza type A, positive antigen test | no | no | no | 2 | 0 |
| 7_B | *Staphylococcus aureus* positive blood culture, femur osteomyelitis on MRI | - | yes | yes | yes | 19 | 3 |

**Supplementary Table S2.** A list of differentially expressed miRNAs comparing bacterial to viral infections.

| **miRNA name** | **Log2FC** | **p-value** | **BH-adjusted p-value** |
| --- | --- | --- | --- |
| ***hsa-miR-136-5p*** | 2,95 | 0,001 | 0,058 |
| ***hsa-miR-514a-5p*** | 2,71 | 0,007 | 0,161 |
| *hsa-miR-514b-3p* | 2,69 | 0,001 | 0,058 |
| *hsa-miR-513a-5p* | 2,52 | 0,004 | 0,128 |
| *hsa-miR-1-3p* | 2,50 | 0,010 | 0,205 |
| *hsa-miR-514b-5p* | 2,20 | 0,014 | 0,231 |
| ***hsa-miR-513c-3p*** | 2,13 | 0,001 | 0,058 |
| ***hsa-miR-514a-3p*** | 1,96 | 0,001 | 0,058 |
| *hsa-miR-513a-3p* | 1,90 | 0,005 | 0,130 |
| ***hsa-miR-507*** | 1,75 | 0,005 | 0,134 |
| *hsa-miR-146b-5p* | 1,73 | 0,037 | 0,433 |
| *hsa-miR-338-3p* | 1,54 | 0,002 | 0,058 |
| *hsa-miR-509-3p* | 1,35 | 0,028 | 0,400 |
| *hsa-miR-181a-5p* | -1,01 | 0,025 | 0,374 |
| *hsa-miR-1180-3p* | -1,10 | 0,044 | 0,433 |
| *hsa-miR-30e-3p* | -1,11 | 0,042 | 0,433 |
| *hsa-miR-652-3p* | -1,12 | 0,039 | 0,231 |
| *hsa-miR-130a-3p* | -1,18 | 0,013 | 0,433 |
| *hsa-miR-203a-5p* | -1,25 | 0,043 | 0,433 |
| *hsa-miR-183-5p* | -1,33 | 0,037 | 0,246 |
| *hsa-miR-30a-3p* | -1,36 | 0,015 | 0,193 |
| *hsa-miR-188-5p* | -1,53 | 0,009 | 0,433 |
| *hsa-miR-888-5p* | -1,75 | 0,040 | 0,433 |
| *hsa-miR-218-5p* | -2,00 | 0,039 | 0,058 |
| *hsa-miR-598-3p* | -2,83 | 0,001 | 0,058 |
| *hsa-miR-542-5p* | -3,74 | 0,002 | 0,374 |

miRNAs were considered differentially expressed if they had a log2FC > 1 or < -1 and a nominal p-value < 0.05. The diagnostic signature to distinguish between infection groups is highlighted in bold.

**Supplementary Table S3.** Confusion matrix and associated internal performance metrics for models for the binary classification of urine miRNA between comparison groups.

| Bacterial vs. Control signature | | | | Viral vs. Control signature | | | | Bacterial vs. Viral signature | | | |
| --- | --- | --- | --- | --- | --- | --- | --- | --- | --- | --- | --- |
|  | | **Predicted** | |  | | **Predicted** | |  | | **Predicted** | |
|  |  | Bacterial | Controls |  |  | Viral | Controls |  |  | Bacterial | Viral |
| Actual | Bacterial | 7 (100%) | 0 (0%) | **Actual** | Viral | 7 (100%) | 0 (0%) | **Actual** | Bacterial | 6 (86%) | 1 (14%) |
|  | Controls | 2 (25%) | 6 (75%) |  | Controls | 2 (25%) | 6 (75%) |  | Viral | 0 (0%) | 7 (100%) |
| Evaluation criteria | | Accuracy | 0.867, (95% CI: 0.595–0.983) | **Evaluation criteria** | | | 0.867 (95% CI: 0.595–0.983) | **Evaluation criteria** | | | 0.929 (95% CI: 0.661–0.998) |
|  |  | Positive prognostic value (PPV) | 0.778 (95% CI: 0.400–0.972) |  |  |  | 0.778 (95% CI: 0.400–0.972) |  |  |  | 1.000 (95% CI: 0.541–1.000) |
|  |  | Matthews correlation coefficient | 0.764 |  |  |  | 0.764 |  |  |  | 0.866 |
|  |  | F1 score | 0.875 (95% CI: 0.584–0.975) |  |  |  | 0.875 (95% CI: 0.584–0.975) |  |  |  | 0.923 (95% CI: 0.621–0.996) |

LASSO regularized regression was used to select diagnostic signatures for distinguishing between comparison groups (bacterial infections vs controls, viral infections vs controls, bacterial vs viral) in febrile children. Evaluation criteria reported with 95% confidence interval where applicable

**Supplementary Table S4.** Overview of selected miRNA potential candidate diagnostic signatures.

| **Name of miRNA** | **Up- or down-regulated (­↑/↓)** | **LASSO coefficients** | **ANOVA F-value** | **ANOVA p-value** |
| --- | --- | --- | --- | --- |
| ***hsa-miR-136-5p*** | ­↑ | 0.768 | 6.80 | 0.010 |
| ***hsa-miR-513c-3p*** | ­↑ | 0.764 | 31.80 | 0.002 |
| ***hsa-miR-514a-5p*** | ­↑ | 0.447 | 7.03 | 0.040 |
| ***hsa-miR-514a-3p*** | ­↑ | 0.404 | 23.27 | 0.010 |
| *hsa-miR-30a-3p* | ↓ | 0.377 | - | - |
| *hsa-miR-888-5p* | ↓ | 0.231 | - | - |
| ***hsa-miR-1-3p*** | ­↑ | 0.160 | 3.72 | 0.019 |
| *hsa-miR-146b-5p* | ­↑ | 0.122 | - | - |
| *hsa-miR-652-3p* | ↓ | 0.062 | - | - |
| ***hsa-miR-507*** | ­↑ | 0.004 | 17.06 | 0.019 |
| *hsa-miR-513a-5p* | ­↑ | - | 9.73 | 0.021 |
| *hsa-miR-542-5p* | ↓ | - | 10.27 | 0.019 |
| *hsa-miR-514b-3p* | ↓ | - | 24.35 | 0.010 |
| *hsa-miR-338-3p* | ­↑ | - | 5.22 | 0.009 |
| *hsa-miR-513a-3p* | ­↑ | - | 29.70 | 0.009 |

The selection process based on LASSO penalized logistic regression between bacterial infections vs. viral infection patients and controls as well as ANOVA for bacterial infections vs. viral infection patients. The miRNA potential candidate diagnostic signature selected for downstream analysis (statistically significant via ANOVA and selected as an important feature by LASSO) is marked in bold.

**Supplementary Table S5.** Internal classification performance and confusion matrix of LASSO regularized regression for the binary classification of blood mRNA between the bacterial infections group and controls.

| **Bacterial vs. Control signature** | | | | **Evaluation criteria** | **Value** | **CI** |
| --- | --- | --- | --- | --- | --- | --- |
|  | | Predicted | | Accuracy | 1.000 | [0.782, 1.000] |
|  |  | Bacterial | Controls | Positive predictive value (PPV) | 1.000 | [0.590, 1.000] |
| **Actual** | Bacterial | 7 (100%) | 0 (0%) | Matthews correlation coefficient | 1.000 | 1.000 |
|  | Controls | 0 (0%) | 8 (100%) | F1 score | 1.000 | [0.646, 1.000] |

LASSO regularized regression was used to select important features for distinguishing between comparison groups (bacterial infections vs controls, viral infections vs controls, bacterial vs viral) in febrile children. The bacterial vs viral comparison did not show sufficient performance and thus not shown, viral vs controls comparisons landed one gene sufficient for comparison so was not evaluated using these metrics.

**Supplementary Table S6.** The results of the Pearson's correlation analysis for miRNA and their respective regulated genes.

| **miRNA name** | **Gene name** | **Pearson’s correlation coefficient** | **p-value** |
| --- | --- | --- | --- |
| *hsa-miR-507* | *ZNF112* | -0.43 | 0.049 |
| *hsa-miR-507* | *TCF7* | -0.45 | 0.035 |
| *hsa-miR-507* | *ABLIM1* | -0.50 | 0.018 |
| *hsa-miR-507* | *BACH2* | -0.59 | 0.004 |
| *hsa-miR-507* | *SLC16A10* | -0.67 | 0.001 |
| *hsa-miR-507* | *ACVR2B* | -0.47 | 0.026 |
| *hsa-miR-507* | *AGMAT* | -0.44 | 0.042 |
| *hsa-miR-507* | *CCR7* | -0.53 | 0.010 |
| *hsa-miR-507* | *MTUS1* | -0.43 | 0.044 |
| *hsa-miR-507* | *PIK3C2B* | -0.46 | 0.031 |
| *hsa-miR-507* | *HRK* | -0.51 | 0.015 |
| *hsa-miR-507* | *RAB30* | -0.46 | 0.030 |
| *hsa-miR-507* | *LEF1* | -0.49 | 0.020 |
| *hsa-miR-507* | *CAMK4* | -0.49 | 0.022 |
| *hsa-miR-507* | *AK5* | -0.57 | 0.006 |
| *hsa-miR-507* | *PLXDC1* | -0.59 | 0.004 |
| *hsa-miR-507* | *IL7R* | -0.51 | 0.015 |
| *hsa-miR-507* | *CNTNAP2* | -0.60 | 0.003 |
| *hsa-miR-507* | *GCNT4* | -0.54 | 0.009 |
| *hsa-miR-507* | *ZNF154* | -0.45 | 0.037 |
| *hsa-miR-507* | *NELL2* | -0.49 | 0.022 |
| *hsa-miR-507* | *PAX5* | -0.43 | 0.047 |
| *hsa-miR-507* | *KLHL14* | -0.45 | 0.038 |
| *hsa-miR-507* | *LBH* | -0.49 | 0.021 |
| *hsa-miR-507* | *SIAH3* | -0.65 | 0.001 |
| *hsa-miR-513c-3p* | *KCNK10* | -0.45 | 0.035 |
| *hsa-miR-513c-3p* | *CUX2* | -0.54 | 0.009 |
| *hsa-miR-513c-3p* | *PASK* | -0.45 | 0.035 |
| *hsa-miR-513c-3p* | *EPHA4* | -0.47 | 0.027 |
| *hsa-miR-513c-3p* | *TNFRSF21* | -0.46 | 0.033 |
| *hsa-miR-513c-3p* | *CDH2* | -0.58 | 0.004 |
| *hsa-miR-514a-3p* | *BACH2* | -0.42 | 0.049 |
| *hsa-miR-514a-3p* | *SLC16A10* | -0.49 | 0.021 |
| *hsa-miR-514a-3p* | *EDAR* | -0.47 | 0.028 |

The analysis was performed between the diagnostic signature of miRNA that distinguishes bacterial and viral infections in febrile children and differentially expressed transcripts among the groups of bacterial and viral infection patients in blood. A statistically significant correlation was defined as having an adjusted p-value < 0.05 and a correlation coefficient < 0.

**Supplementary Table S7.** A list of miRNAs that are differentially expressed in the bacterial or viral patient group compared to the controls, but not differentially expressed when comparing the patient groups to each other.

| **miRNA name** | **Up- or down-regulated in B vs C group (­↑/↓)** | **Up- or down-regulated in V vs C group (­↑/↓)** |
| --- | --- | --- |
| *hsa-let-7f-1-3p* | ­↑ | ­↑ |
| *hsa-miR-12136* | ↓ | ↓ |
| *hsa-miR-1246* | ↑ | ↑ |
| *hsa-miR-1277-5p* | ↑ | ↑ |
| *hsa-miR-1290* | ↑ | ↑ |
| *hsa-miR-200c-3p* | ↓ | ↓ |
| *hsa-miR-204-3p* | ↑ | ↑ |
| *hsa-miR-22-5p* | ↑ | ↑ |
| *hsa-miR-25-3p* | ↓ | ↓ |
| *hsa-miR-27b-5p* | ↑ | ↑ |
| *hsa-miR-3065-3p* | ↑ | ↑ |
| *hsa-miR-3065-5p* | ↑ | ↑ |
| *hsa-miR-375-3p* | ↓ | ↓ |
| *hsa-miR-451a* | ↑ | ↑ |
| *hsa-miR-502-3p* | ↑ | ↑ |
| *hsa-miR-508-3p* | ↑ | ↑ |
| *hsa-miR-508-5p* | ↑ | ↑ |
| *hsa-miR-510-5p* | ↑ | ↑ |
| *hsa-miR-545-5p* | ↑ | ↑ |
| *hsa-miR-708-3p* | ↑ | ↑ |
| *hsa-miR-99b-5p* | ↓ | ↓ |

**Supplementary Figure S1.** Selection of miRNA binary signatures from urine to distinguish infection etiology in febrile children.

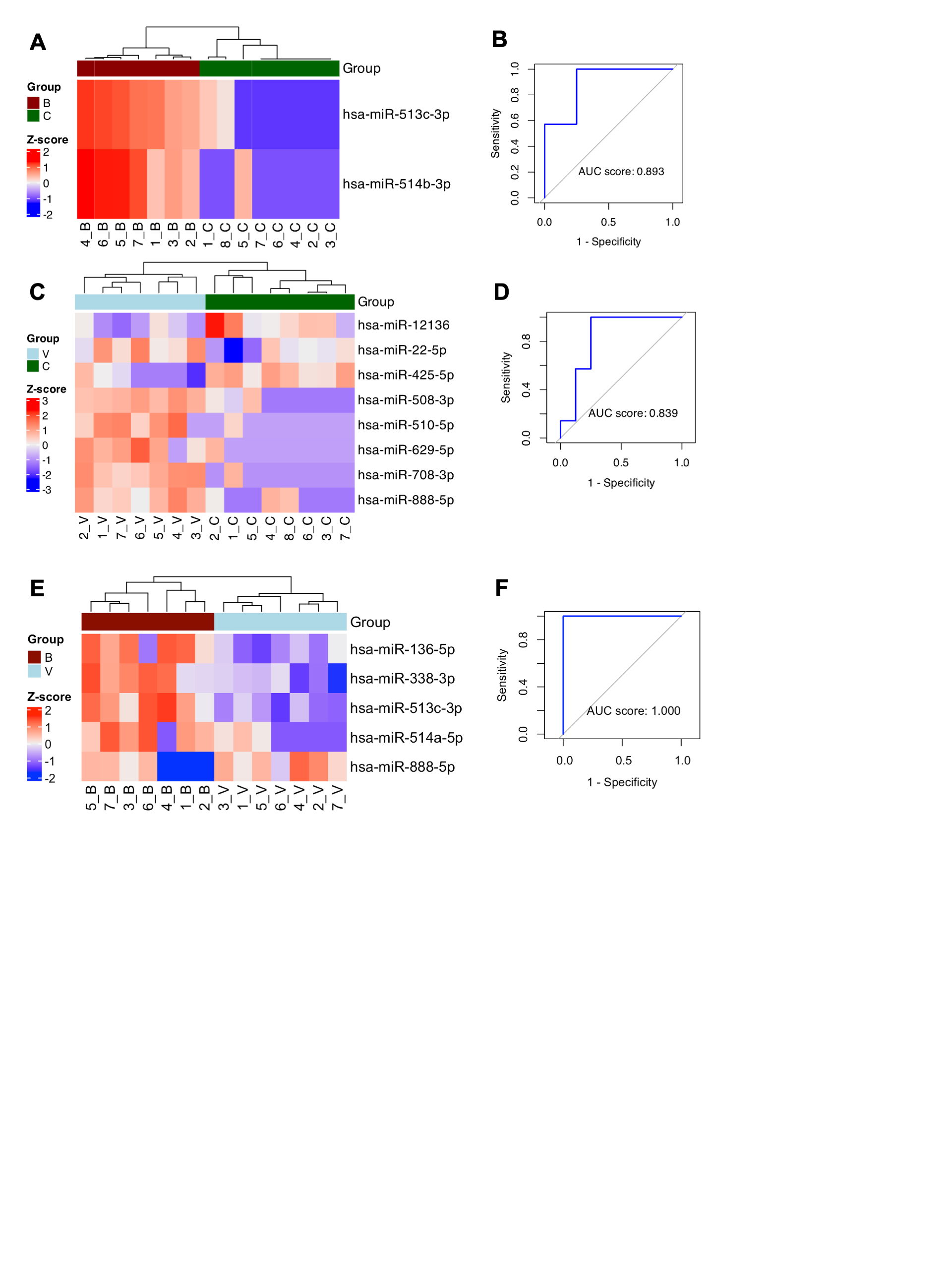

The analysis was performed using binary LASSO logistic regressions that compare the group of bacterial infections (n = 7) vs. controls, the group of viral infections (n = 7) vs. controls, and the group of bacterial vs. viral infections.

Heatmaps along with agglomerative hierarchical cluster analysis using Z-score for selected miRNA biomarker candidates that differentiate between bacterial infections and controls (A), viral infections and controls (C) and bacterial and viral infections (E). Negative Z-score marked in blue, positive Z-score marked in red, Z-score reaching zero marked in white. In cluster analysis patients with bacterial infections marked in dark red, with viral infections – in light blue, controls – in green. ROC curves for bacterial infections vs controls (B), viral infections vs controls (D), and bacterial vs viral infections (F) reveal AUC scores of 0.893, 0.839 and 1.000 respectively.

**Supplementary Figure S2.** ROC curve for model used for miRNA selection.

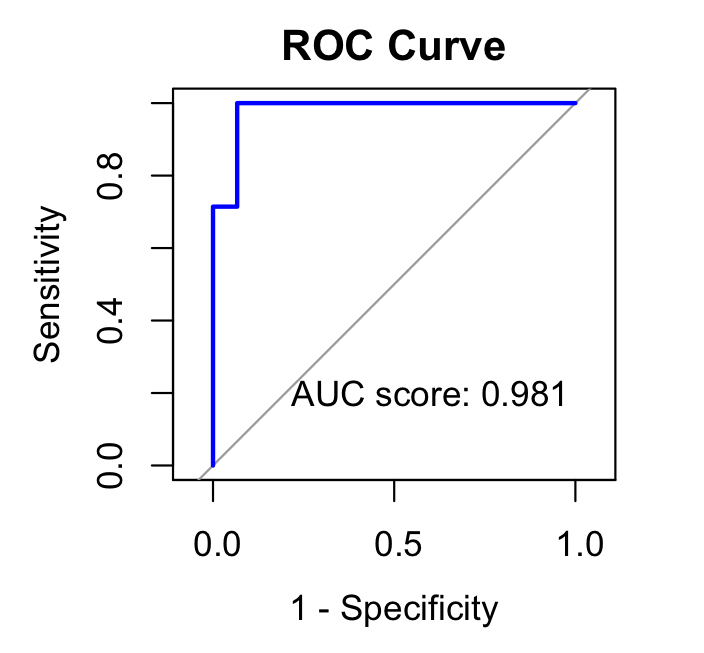

The LASSO regularized regression was implemented to prioritize urine miRNA signature to distinguish bacterial and viral infections in febrile children. The diagonal line represents random predictions, and blue represents the empirical ROC curve. False positive rate or sensitivity marked on the x-axis, true positive rate or 1-specificity marked on the y-axis.

**Supplementary Figure S3.** Selection of mRNA blood-based binary signatures to distinguish infection etiology in febrile children.

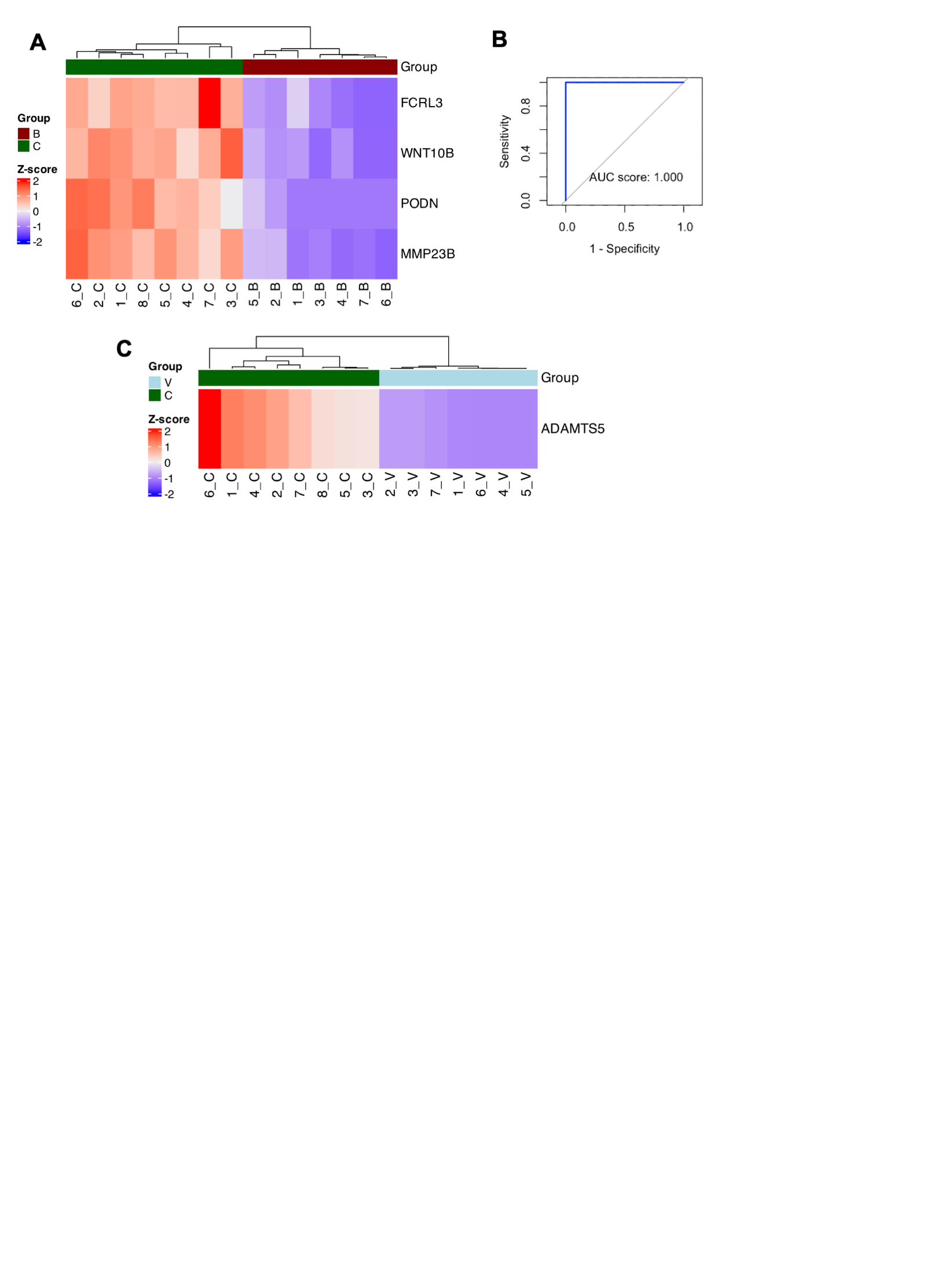

LASSO regularized regression was used that compare the bacterial infection group (n=7) vs controls and the viral infection group (n=7) vs controls. Heatmaps along with agglomerative hierarchical cluster analysis using Z-score for selected mRNA signature differentiating between bacterial infections and controls (A) and viral infections and controls (C). Negative Z-score marked in blue, positive Z-score marked in red, Z-score reaching zero marked in white. In cluster analysis, patients with bacterial infections marked in dark red, with viral infections – in light blue, controls – in green. The ROC curve for bacterial infections vs. controls (B) reveals an AUC score of 1.000.
